# In the pursuit of longevity: anti-aging substances, nanotechnological preparations, and emerging approaches

**DOI:** 10.1101/2022.03.20.22272670

**Authors:** Elena Giannouli, Vangelis Karalis

## Abstract

**Objectives:** To summarize information on anti-aging substances and nanotechnology formulations against aging and to unveil emerging anti-aging approaches like gene therapy, stem cells, monoclonal antibodies, and future trends.

**Methods:** A systematic literature search was conducted through May 10, 2021. Eligibility criteria were articles in English relevant to the aim of this review.

**Results:** Anti-aging substances like coenzyme Q10, curcumin, ginsenoside Rg1, bioidentical hormones, and geroprotectants, can help to slow the aging process. Nano-delivery of anti-aging substances are novel approaches able to delay aging. Cryogenic sleep, therapeutic hypothermia, and caloric restriction are promising interventions that may one day grant humans biological immortality.

**Discussion:** The ability to target basic maturation mechanisms, such as responses to oxidative damage, inflammation, irritation, and senescence associated with multiple concurrent deficits, is the most important feature. Nanocarriers to transport bioactive compounds have been shown to improve their stability, increase intestinal absorption, and prolong their circulation time.

## Introduction

All living organisms undergo systemic physiological deterioration after ontogeny, which is referred to as aging [1]. A crucial first step in the field of aging research was an observation in 1939 where caloric restriction increased the lifespan of mice and rats. This was the first demonstration of the plasticity of the aging process and was a foreshadowing of the genetic studies that would follow 50 years later. In the 1900s, scientists began to consider whether aging was the cause of age-related chronic diseases [1]. Many of the molecular and biochemical mechanisms that determined the rate of aging were also being studied in laboratories that focused on specific chronic diseases. Because different species have radically different lifespans, ranging from days to decades, biologists have long recognized lifespan as a heritable trait with a genetic basis [2]. Medawar proposed in 1952 that aging is caused by a decrease in the power of natural selection [2]. Over the past 30 years, the focus of aging research has shifted from the identification of aging phenotypes to the study of the genetic pathways that determine these phenotypes. A complex network of interconnected intracellular signaling pathways and higher order processes has been discovered by aging genetics. Many of the identified pathways and cycles are known to play a role in homeostatic responses to environmental changes [2]. Each of these lines of research adds a crucial piece to the complexity puzzle of aging, and it will be critical to integrate them over time. Although there is no single “key” to understanding aging, these studies have shown that the rate of aging can be slowed, implying that managing aging will also, coincidentally, slow the onset of or reduce the burden of many diseases and extend life expectancy. Maturation in daily life can begin at a very young age, according to some findings. The energy requirements and pressures of recombination, as well as the death of each generation, benefit the evolutionary process. This may lead us to believe that maturation is an adaptive and even an altruistic program in which every cell and organism that has ever existed eventually dies for the survival and benefit of the entire biological unit [3].

Aging is associated with the accumulation of deleterious changes (from cell senescence to a decrease in immune function and hormone secretion) over time, resulting in a loss of function in multiple tissues and an increase in the likelihood of death. Changes in gene expression that occur normally over the lifetime of an organism without adjustment of DNA arrangement are referred to as: “aging epigenetics” [4]. Multiple enzymes regulate chromatin epigenetics, resulting in changes in DNA methylation and histone methylation/acetylation events. Aneuploidy is characterized as an unusual chromosome number caused by mis-segregation of chromosomes during cell division in both germ cells and somatic cells. It has been associated with cellular senescence and early maturation in mice, supporting its possible role in age-related loss of tissue homeostasis. Genomic instability, telomere decay, epigenetic drift and deficient proteostasis have all been shown to influence mitotic disfigurement and aneuploidization [4].

Efforts and attempts to control aging have been important since early developments in human culture. In the 3^rd^ century BC, Chinese Taoists invented a system of measures to prolong life. Today, perhaps more than ever, anti-aging research is a global focus. The aging of citizens is emerging as an undeniably significant financial and social challenge for all nations [5].

The aim of this review is to summarize the available information on anti-aging substances and nanotechnology formulations and to present novel emerging anti-aging approaches such as gene therapy, the use of stem cells and monoclonal antibodies for anti-aging purposes, and trends that will rise in the near future.

## Methods

### C1. Search strategy

A systematic search of the PubMed database was conducted through May 10, 2021. The terms searched were “anti-aging” + “substances”, “nanotechnology formulations” + “aging”, “gene therapy” + “anti-aging”, “stem cells” + “anti-aging treatment” + “rejuvenation”, “monoclonal antibodies” + “anti-aging”, “anti-aging” + “future prospects”. Eligibility criteria were: Articles written in English relevant to the aim of this review. The results of the search strategy are shown in Table 1.

**Table 1.** Search terms and number of citations retrieved.

| **ANTI-AGING STRATEGIES** | **SEARCH STRATEGY** | **CITATIONS RETRIEVED (26.03.2021)** | **STUDIES RETRIEVED AFTER SCREENING OF TITLE AND ABSTRACT** |
| --- | --- | --- | --- |
| “ANTI-AGING” + “SUBSTANCES” | PubMed | 8,448 | 42 |
| “NANOTECHNOLOGICAL FORMULATIONS” + “AGING” | PubMed | 286 | 25 |
| “GENE THERAPY” + “ANTI-AGING” | PubMed | 880 | 38 |
| “STEM CELLS” + “ANTI-AGING TREATMENT” + “REJUVENATION” | PubMed | 571 | 16 |
| “MONOCLONAL ANTIBODIES” + “ANTI-AGING” | PubMed | 157 | 5 |
| “ANTI-AGING” + “FUTURE PERSPECTIVES” | PubMed | 761 | 22 |

### C2. Selection criteria

After removing duplicate articles, all studies were evaluated using the following criteria: a) journal, b) authorship, c) publication date, d) study design, e) methods of analysis, f) results, and g) conclusions. Selection criteria were: articles in English and relevance to the aim of this review. To improve data quality, all studies that met the inclusion criteria were thoroughly assessed for rationale, method design, primary outcome, fatigue assessment, statistical analysis, results, discussion, and conclusions. Those studies that had any bias in methodology, results, or interpretation of exposed data that could be reflected in the overall study analysis were also excluded.

## Results

### Anti-aging substances

#### Bioidentical hormones

Hormone replacement therapy (HRT) within the United States experienced significant growth and acceptance during the 1980s and -90s, peaking in 1999 with well over 85 million medications prescribed for women alone [6]. Since then, “anti-aging” specialists have heralded another, more protected and seductive option: bioidentical hormone replacement therapy (BHRT). BHRT was sweepingly recommended in self-improvement guidebooks, in the mainstream media, and by TV or social media superstars as a panacea for maturation and menopause. The allure and guarantee of BHRT lies in the belief that it is a “natural” treatment and therefore poses fewer dangers than artificial hormones, while offering menopausal relief and surprisingly anti-aging benefits [6].

#### Geroprotectors

Advances in the field of aging research have illuminated the way for the blossoming of drugs with the ability to target weakness, commonly referred to as “geroprotectors” [7]. The most important feature for this class of drugs is the ability to target basic mechanisms of aging, such as responses to oxidative damage, inflammation, irritation, senescence, which are associated with various co-occurring deficits, such as hearing loss, signs of tremor, dementia, which may occur with aging [8]. It is considered that they may be able to delay or even reverse the frailty of the elderly and also improve their responses to unfortunate circumstances.

Geroprotectors consist of more than 200 substances, each of which is believed to extend the lifespan of organisms [9]. Geroprotectors have shown the ability to delay the onset of dysregulations in various tissues that lead to age-related diseases by modifying aging processes such as autophagy, senescence, and inflammation [8, 10]. Many compounds such as rapamycin, resveratrol, metformin and senolytics such as fisetin, dasatininb, quercetin that inhibit cell proliferation can prolong life span in various organisms.

Many expectations are placed on the use of senotherapeutics, especially senolytics that remove senescent cells and senostathics that kill the SASP (senescence-related secretory phenotype) [11]. Aggregation of senescent cells has been shown to be responsible for weakness and age-related diseases [12]. A plethora of senotherapeutics is repurposed and well-studied drugs. To limit side effects, senolytics should be administered in sporadic doses. Studies with quercetin together with dasatininb and fisetin have performed under different conditions and with healthy elderly participants [13].

#### Coenzyme Q10

Coenzyme Q (CoQ), ubiquinone, or 2,3-dimethoxy-5-methyl-6-polyprenyl-1,4-benzoquinone is a bipartite particle. CoQ in eukaryotes occurs primarily in mitochondria through a group of nuclear-encoded CoQ proteins via a biochemical pathway that is not yet fully defined [14]. The ability of CoQ to act as a cancer preventive agent may be important in maturation, as it is commonly reported that oxidative stress increases with aging. However, the lack of longevity effects in many studies means that these conditions should not be relied upon exclusively [14].

#### Curcumin

Curcumin, a polyphenolic compound extracted from *Curcuma longa*, exhibits potent anti-aging properties. Recently, research on curcumin in relation to maturation and age-related diseases in organic model organisms has shown that curcumin and its metabolites have prolonged the mean lifespan of some aging model organisms, such as *C. elegans, D. melanogaster*, yeast, and mouse. In addition, curcumin is involved in many biological activities such as antioxidant, anti-inflammatory, anti-cancer, chemopreventive, and against neurodegenerative traits [15].

Several studies on different organisms have shown that prolonged life span with the use of curcumin is associated with increased superoxide dismutase and decreased levels of malondialdehyde and lipofuscin. Also, the crucial role of curcumin in setting important markers that affect the age of organisms such as markers IIS, mTOR, PKA, FOXO [14]. It was demonstrated that brain tumor cells showed a decrease in telomere length after long-term treatment with curcumin. Similarly, it was found that addition of curcumin resulted in enhancement of deleterious effects on DNA and then on transcription factors of brain tumor cells. Long-term administration of curcumin improved motor function in moderately aged rhesus monkeys. Curcumin supplements additionally improved motor function and also cognitive function in healthy middle-aged adults [16]. Curcumin in reducing cell irritation related to neurodegenerative and maturation procedures is associated with extended CISD2 articulation.

#### Ginsenoside Rg1

Ginseng is a major Chinese indigenous drug known by its blood nourishing role as described by the traditional Chinese medicine hypothesis. The advanced clinical hypothesis [17] additionally supports that ginseng is characterized by the anti-inflammatory, anti-aging, anti-oxidant, anti-injury and also immunity-boosting qualities.

Additionally, announced that Rg1 could prolong the existence of mice and postpone the maturation of human lung fibroblasts. Rg1 was known to slow down tert-butyl hydroperoxide-activated maturation of HSCs (Hematopoietic Stem Cells) through signaling pathways regulating p19arf, p16INK4a, p21Cip/Waf1, and p53. The mechanism that slows the maturation of HSC/HPC cells is uncertain; focusing on molecules that affect maturation postponed by Rg1 is currently an important matter for further research [18]. In addition, Rg1 was shown to protect the hippocampus from irregularities in a rodent model of maturation triggered by D-galactose treatment.

ROS (Reactive Oxygen Species) aggregation induced aging and age-related disorders. Long-term treatment with D-gal can induce maturation symptoms such as intellectual disorders, shortened life expectancy, oxidative stress, neurodegeneration, and immunological defects [19]. Researchers discovered that Rg1 treatment surprisingly decreased the percentage of G0/G1 stage cells and SA-b-Gal-positive staining Sca-1+ HSC/HPC cells and significantly expanded CFU (Colony Forming Unit) mix values in contrast to the D-galactose model. All these results suggest that Rg1 treatment has an anti-aging effect on Sca-1+ HSC/HPC cells and drives cell expansion in the maturing rodent model [19].

#### Seaweed derivatives

The life-prolonging effects of substances from marine macroalgae have been studied in animal models, such as yeasts and flies. Fucoxanthin, a carotenoid and a major photosynthetic shade derived from brown algae, showed an extension of life expectancy in both *Drosophila melanogaster* and *Caenorhabditis elegans* [20]. These findings suggest that porphyran can extend life expectancy and health span [21].

Similarly, polysaccharides from the brown alga *Saccharina japonica* can remarkably increase normal life expectancy in both male and female *Drosophila*. A wealth of data regarding algal substances and their anti-cellular senescence effects is known. *Fucoidan* is an algal compound that preserves cells from both replicative and stress-induced senescence. discovered that fucoidan decreased senescence in cultured colony-forming endothelial cells with long residence time, as demonstrated by attenuation of senescence-induced beta-galactosidase (SA -β-Gal) activity [22].

The SA -β-Gal activity is the most commonly used biomarker to detect senescent cells. Another study has described that fucoidan treatment decreases SA-β-Gal activity initiated by p-cresol, a major uremic toxin, in mesenchymal stem cells [22]. The anti-senescence role and life-prolonging effects of algal derivatives in lower life forms support research on the anti-aging effects of algal compounds in higher organisms. Despite the fact that the effects of algal derivatives on key signaling pathways controlling life expectancy have been described, the life-prolonging effect of algal-derived bioactive compounds has not yet been assessed using mammalian models [22].

#### Leontopodium alpinum callus culture extract

The efficacy of edelweiss callus culture extract (*Leontopodium Alpinum* callus culture extract, LACCE) was investigated using various assays from in vitro to in vivo as well as transcriptome profiling. Some in vitro assay results indicated the important antioxidant properties of LACCE in relation to UVB treatment. In vivo clinical test showed that continuous application of LACCE on the face and skin tissues improved wrinkling, skin elasticity, skin density and skin thickness in contrast to placebo treatment. RNA sequencing results showed that at least 16.56% of human genes were expressed in keratinocyte cells [23].

The genes upregulated by LACCE encoding some KRT (Keratin) proteins, DDIT4, BNIP3 and IGFBP3, were involved in the positive regulation of the developmental process, programmed cell death, keratinization and keratinization, resulting in skin barriers that provide numerous benefits to human skin. On the other hand, the down-regulated genes were stress-responsive genes, including metal, oxidation, injury, hypoxia, and viral infection, suggesting that LACCE does not induce negative effects, such as stress, on the skin. The whole study showed that LACCE is a potential and very effective agent for anti-aging cosmetics [23].

### Nanotechnological formulations against aging

#### Nano systems

Nanosystems represent a promising platform that can circumvent a variety of difficulties associated with the conventional treatment of neurological disorders, such as nanoemulsions, dendrimers, polymeric nanoparticles, and carbon nanotubes. Although tremendous progress has been made in the field of neuro-nanomedicine over the past decade, the clinical fraternity remains hamstrung by the inadequate development of nanosystems from benchtop research facilities to clinical trials and applications. Regulatory agencies in combination with industry must essentially address the lack of regulatory requirements for large scale nanosystems manufacturing and clinical trials. The development of a concise and comprehensive framework for nanosystems must define the fate of nanosystems, the issues of toxicity, long-term exposure, and also the potential environmental impacts [24].

#### Nanoemulsions

Nanoemulsions are characterized as a kinetically stable biphasic colloidal dispersion containing two immiscible liquids and an emulsifier. They exhibit enticing and helpful properties such as adjustable rheology, optically transparent appearance and increased surface area per unit volume. A risperidone-loaded nanoemulsion with water and an emulsifier increases the uptake of the active drug moiety by 1.3 times compared to a risperidone solution. This nanosystem is an attractive drug delivery vehicle for targeted delivery to the central nervous system [25].

#### Dendrimers

Dendrimers are three-dimensional, hyperbranched, multivalent, spherical polymeric nanocarriers that exist within the particle diameter of 1-10 nm. The well-characterized spherical architecture of dendrimers confers monodispersity and propensity for functional changes. Swami and co-workers found higher drug uptake compared to unconjugated nanodendrimers or conventional commercial drugs [26]. Other use cases are not included in Table 1.

#### Polymeric nanoparticles

Polymer-based nanocarriers provide an effective and adaptable platform that allows nanoparticles to be modified to meet specific criteria. These properties include higher drug encapsulation efficacy, surface conformability, ease of fabrication, and safety against degrading enzymes and other chemicals. The appealing multilayer properties of natural or manufactured polymers make them ideal candidates as drug carriers for nanoparticles [27].

#### Carbon nanotubes

Carbon nanotubes (CNTs) are the allotropic form of carbon that belongs to the fullerene family. CNTs exist as elongated and cylindrical molecules that conform to a hexagonal arrangement of hybridized carbon atoms. Drug delivery is achieved by consolidating the ideal molecule on the surface or tip of the CNT. Nanosystems represent a promising platform that can circumvent a variety of difficulties in the conventional treatment of neurological diseases [28].

#### Nanodelivery of phytobioactive compounds

Phytonanotherapy is a promising technique that can overcome some of the problems present in standard therapy directions. Nanocarriers for transporting phytobioactive compounds (PBCs) improve their stability and solubility, increase their absorption in the gastrointestinal tract, protect against premature enzymatic degradation and absorption, prolong their circulation time, and thereby limit the negative side effects of these compounds. The nanocarrier could be clinically similar to standard treatment with synthetic drugs, but with much fewer disadvantages [29]. This would provide an alternative option to standard therapeutic methods for the treatment of age-related diseases.

#### Nano-phytoantioxidants

Nano-phytoantioxidants have the potential to prevent and treat a variety of age-related pathological conditions. In some animal models, orally administered nano-phytoantioxidant nanoparticles showed more impressive potential in combating cardio-metabolic diseases than their natural forms. For example, significant antidiabetic potential was shown in diabetic mice administered nanoparticles loaded with the isoquinoline alkaloid berberine. These mice showed considerable retention of body weight gain and also improved glucose elasticity and insulin sensitivity [30]. Nanotechnologically engineered phytobioactive substances such as resveratrol, curcumin, berberine, epigallocatechin gallate (EGCG), aloe-emodin, and oridonin, exert antioxidant and anti-inflammatory activities. Nanocomposites loaded with these compounds showed enhanced anti-tumor activity compared to unmodified compounds. The efficacy of nanotechnology-based systems has also been demonstrated in combating neurodegenerative diseases such as Alzheimer’s and Parkinson’s disease. The development of such methods seems to be of particular importance for this therapeutic area, as the treatment of these diseases is quite a difficult task due to the blood-brain barrier. Nanotechnological methods also seem to have therapeutic potential in the treatment of Parkinson’s disease. Nanobiotechnology-based nanoparticles loaded with ascorbic acid exert antioxidant and anti-inflammatory activities, have been shown to enhance aggregation of EGCG in the cerebrum, increase the number of synapses, and decrease amyloid-beta plaque/peptide aggregation and neuroinflammation. In a mouse model of familial Alzheimer’s disease, oral nanoparticle administration improved EGCG aggregation, expanded synapses, and decreased amylidotransferrin levels [31].

#### Promotion of the stability of lycopene

Overproduction of reactive oxygen species (ROS) causes great damage to cell structure and functionality. Antioxidants are compounds that can prevent, eliminate, or delay the oxidation of certain molecules. The use of antioxidants in medicine has limited digestibility, instability, inability to pass membrane barriers, and rapid cell release. Liposomal delivery systems are promising candidates for biocompatible, biodegradable, non-toxic synthesized phospholipid vesicles. Lycopene in liposomal formulations could be of great benefit in the treatment of numerous diseases in which oxidative stress plays a critical role. Liposomes are exceptionally effective in delivering antioxidants and achieving prophylactic and therapeutic effects against aging [32].

#### Layer by layer functionalization for oral liposomal formulations

Layer by layer (LbL) functionalized liposomes are prepared in three steps: preparation of the liposome core, LbL coating and further surface modifications if required. LbL technology has proven to be ideal for the production of new drug formulations for a wide range of therapeutic agents. However, the improvement of LbL innovation for oral delivery is still in its early stages with several technical challenges, such as concerns about toxicity, high polydispersity and particle aggregation [33].

### Novel anti-aging approaches

#### Gene therapy

Klotho (Kl) is an aging silencer gene. It has been described that compound H initiates Kl shaping, but its mechanism of induction has never been studied. The effect of compound H in inducing KL articulation was much stronger than that of 1,25-dihydroxy vitamin D (calcitriol), an agonistic ligand for vitamin D receptors known to initiate KL expression. This suggests that induction of Kl expression may be a successful therapeutic technique to prevent or ameliorate age-related diseases. However, it remains a challenge due to the lack of information on the induction mechanism [34]. Using gene knockouts, researchers genetically engineered mice to lack the CISD2 gene. Mice without that gene showed signs of premature aging, such as cell death and degeneration of neurons and muscle cells [35]. Another option relies on the sirtuin family of genes. The sirtuin genes are a family of seven genes that code for proteins found in the evolution of numerous life forms. Expression of these genes appears to play an important role in age-related medical problems such as cancer and deregulated metabolism. Mice lacking SIRT6 age faster, with signs of degeneration, suggesting that SIRT 6 may be important for healthy DNA repair [36].

Telomerase activation is presented as a probable technique to restore tissues and treat diseases represented by premature telomere shortening [37]. Gene therapy approaches are nowadays considered as a way to introduce genes into adult tissues to repair genetic defects or diseases. It has been shown that expanded TERT (telomerase reverse transcriptase) expression in adult and aged mice by using a gene treatment method has regenerative effects without increasing cancer risk [37].

#### Stem cells

Stem cells act as an organizing framework for the body, regenerating and replacing aging cells to keep the entire organism healthy. Mesenchymal stem cells can affect white blood cell processes. They give the human body more tools to fight the natural aging process by remarkably reducing inflammation [38].

Platelet Rich Plasma (PRP) is an exceptionally effective cell product derived exclusively from our own blood. It contains numerous beneficial growth factors that have proven effective in a variety of applications, including orthopedics, cosmetics and wound healing. PRP is typically derived from 20-40 ml of the patient’s own blood and then re-injected where needed, or combined with stem cells whenever necessary. Common orthopedic conditions for which PRP can be used include osteoarthritis of the knee, hip, spine and shoulder, tennis elbow, Achilles’ tendonitis etc. [39].

Adipose-derived Stem Cells (ASCs) have some advantages in clinical applications as they are not difficult to obtain and are abundant in the human body, which means that there are no moral problems in obtaining these cells. The anti-aging effects of ASCs have been investigated, particularly the suppression of the glycation response and the restoration of skin functionality in a mouse model of accelerated aging induced by D-Gal [40].

#### Monoclonal antibodies

Advanced glycation end (AGEs) products are implicated in several age-related diseases and conditions, e.g., diabetes mellitus (DM) and its complications, Alzheimer’s disease, vascular dementia, atherosclerosis, hypertension, and chronic kidney disease. The presence of AGE - modified IgG may also have a defensive role and serve as part of the mechanism to clear damaged or dysfunctional proteins. However, their existence has been confirmed by some studies showing that these anti-AGE -Abs are present in healthy individuals as well as in various diseases [41].

## Discussion

### Overall

Over time, aging is associated with a cascade of negative changes (from cell senescence to a decrease in immune function and hormone secretion). Age epigenetics refers to changes in gene expression that occur naturally during an organism’s lifetime without affecting DNA arrangement. Aneuploidy is defined as an abnormal chromosome number caused by chromosome maldistribution during cell division in both germ cells and somatic cells. It has been shown to be capable of driving progression to a fully senescent state.

Autophagy, also known as “self-consumption,” is an important target in biomedical research. In a variety of model systems, acetyl-CoA regulates lifespan and autophagy. This suggests that it may act as a sensor, integrator, and transducer for both the overwhelming effects of food intake and the defensive effects of CR [3]. As an organic unit ages, it accumulates more fat and its ability to induce autophagy decreases. Although aging is associated with a decrease in the ability to respond to stress and an increase in the incidence of pathology, death remains the ultimate consequence of aging [42]. According to Hayflick, one of the leading causes of death in old age could be cured. Recent evidence suggests that autophagy as well as anti-aging substances such as coenzyme Q10, curcumin, ginsenoside Rg1, bioidentical hormones, and geroprotectants may help slow the aging process. The ability to target basic maturation mechanisms, such as responses to oxidative damage, inflammation, irritation, and senescence associated with multiple concurrent deficits, such as hearing loss, tremors, and dementia that can occur with aging [43], is the most important feature for this new class of drugs. Therefore, scientists believe that they may be able to delay or even reverse the onset of frailty in the elderly, as well as improve their responses to adversity. More than 200 substances make up the geroprotectors, each of which is believed to help organisms live longer [44].

Curcumin, resveratrol, genistein, and quercetin are examples of phytochemicals thought to have great potential for preventing and treating age-related chronic oxidative stress and systemic inflammation [29]. Nevertheless, their stability in the gastrointestinal tract (GI), bioavailability and solubility have so far limited their clinical application. The use of nanocarriers to transport phytobioactive compounds has been shown to improve their stability and solubility, increase GI absorption, protect against premature enzymatic deterioration and absorption, and prolong their circulation time, thus limiting the negative side effects of these compounds. In this context, the efficacy of nanotechnology formulations in the fight against aging was also evaluated [29]. Nanodelivery of phytobioactive compounds, liposomal formulations that promote the stability of lycopene, triple nanoemulsions with microencapsulated retinol, heptapeptide-loaded solid lipid nanoparticles, and injection of hyaluronic acid are just some of the novel anti-aging approaches being explored.

In addition, gene therapy targeting the anti-aging gene Klotho, APOE alleles, sirtuin genes, and telomerase gene therapy could slow physiological aging and extend life expectancy by about 20% [45]. Moreover, stem cells and their ability to express telomerase, rejuvenate themselves, and differentiate into all three embryonic tissues pose a threat to aging as a process that is not inevitable and can even be reversed. By replacing apoptotic and necrotic cells with healthy ones, stem cells can help regenerate aged tissues and organs. Monoclonal antibodies and anti-aging clinical trials such as reprogramming, senolysis and cell killing strategies, on the other hand, have shown promising success in combating aging and delaying age-related diseases [46].

### Future perspectives

Cryosleep is a process in which an individual is put into a state of suspended animation using medication or a chamber or something very cold, and it is a typical science fiction trope. It might be known as a way to cheat death, whereby a human’s life form can be suspended at -200 degrees Celsius and most likely preserved in liquid nitrogen. The problem with cryogenics and cryosleep is pure physics: our cells are filled with water and when water freezes, it expands and forms crystals, irreversibly damaging the body [47].

The gut microbiome has been proposed as a potential determinant of healthy aging. Undoubtedly, maintaining homeostasis between host and microbiome can prevent inflammation, gut permeability, and decline in bone and cognitive health [48]. For the first time, a phylogenetic microbiota analysis of semi-supercentenarians, i.e., from 105-109 years of age, compared to adults, elderly and centenarians was performed, and as a result, the longest available human microbiota trajectory along aging was reconstructed.

Temperatures have profound effects on behavior and aging in poikilotherms and homeotherms. The longevity of worms, flies and fishes shows an inverse association with ambient temperature. This could be due to the fact that chemosensory neurons modulate life expectancy by likely detecting chemical cues from the environment. For example, when the core body temperature of mice is lowered, life expectancy increases. Lower body temperature is associated with longer life expectancy in both mammals and humans [49]. Caloric restriction (CR) without malnutrition delays aging and extends life expectancy in model organisms and rodents. CR exerts its anti-aging effects via small noncoding RNA molecules, diet composition, and also IGF-1 signaling [50].

## Conclusion

Aging is a natural, heterogeneous feature of most living organisms, especially mammals, and current research has provided clues on how to improve some converging cellular processes related to nutrient recognition, mitochondrial bioenergetics, and healthy senescence. Anti-aging substances such as coenzyme Q10, curcumin, ginsenoside Rg1, bioidentical hormones, and geroprotectors can help slow the aging process. Nano-delivery of anti-aging substances is a novel approach to delaying the onset of aging. Cryogenic sleep, therapeutic hypothermia, and caloric restriction are promising interventions that could grant humans biological immortality one day. The most important feature is the ability to target basic maturation mechanisms, such as responses to oxidative damage, inflammation, irritation, and senescence associated with multiple concurrent deficits. It has been demonstrated that using nanocarriers to transport bioactive compounds improves their stability, increases intestinal absorption, and extends their circulation time.

## Data Availability

All data produced in the present work are contained in the manuscript

## Figures

**Figure 1.**
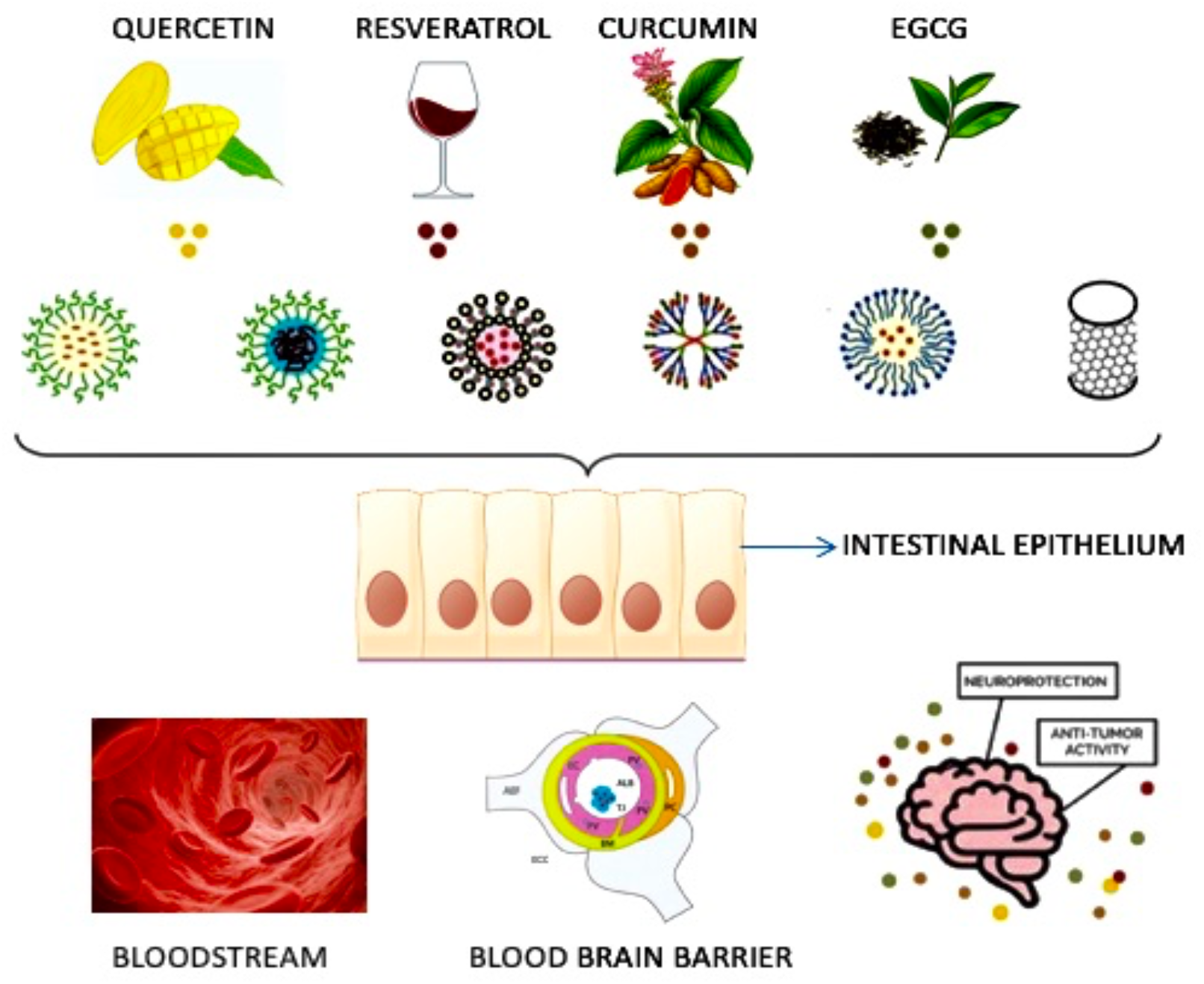
Schematic representation of nanotechnology-based systems used to deliver phytoantioxidant-loaded nanodelivery systems to the brain (adapted from [29]).

**Figure 2.**
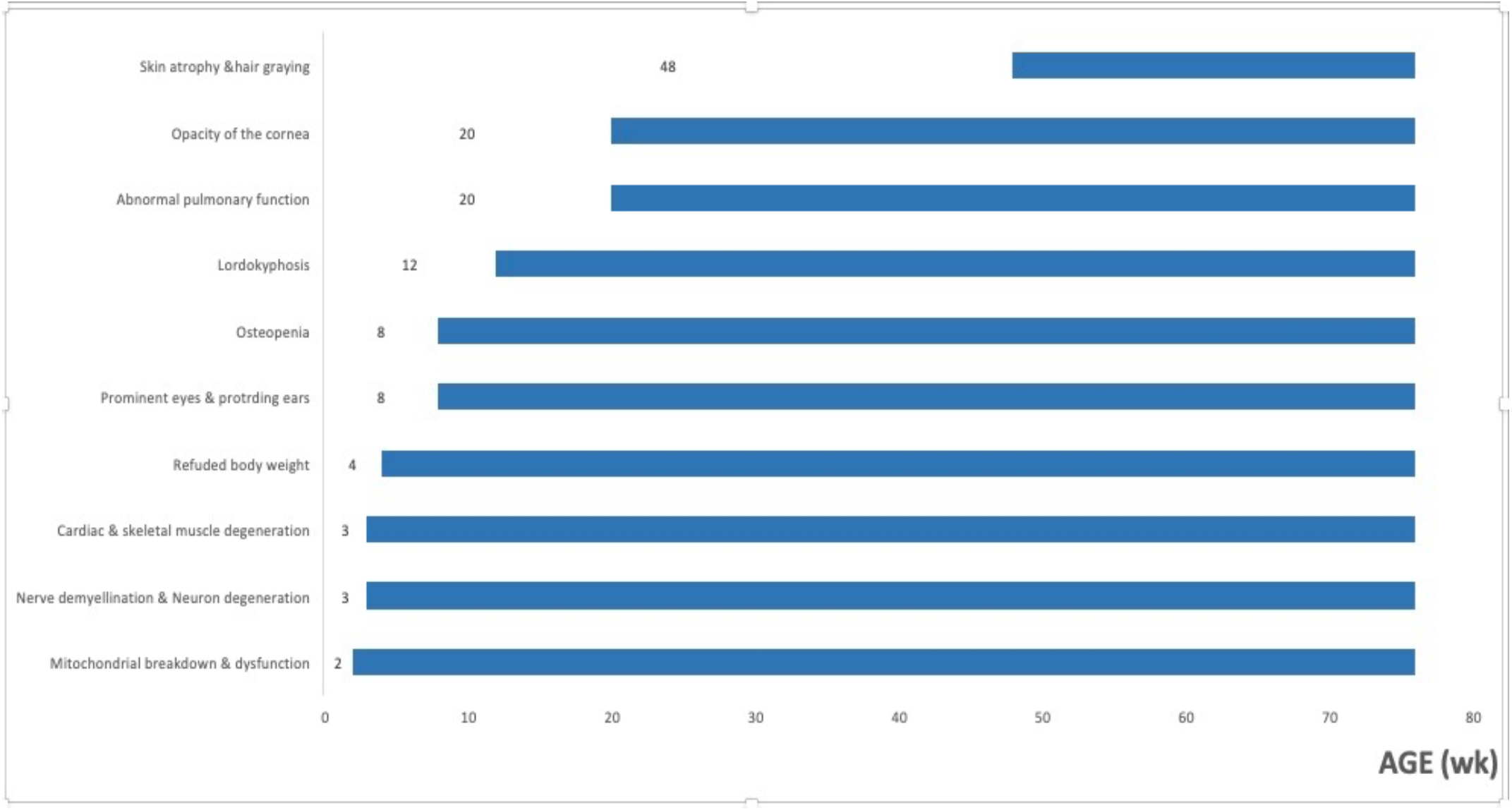
Summary of aging-related phenotypes as a function of age in Cisd2 mice (adapted from [35]).

**Figure 3.**
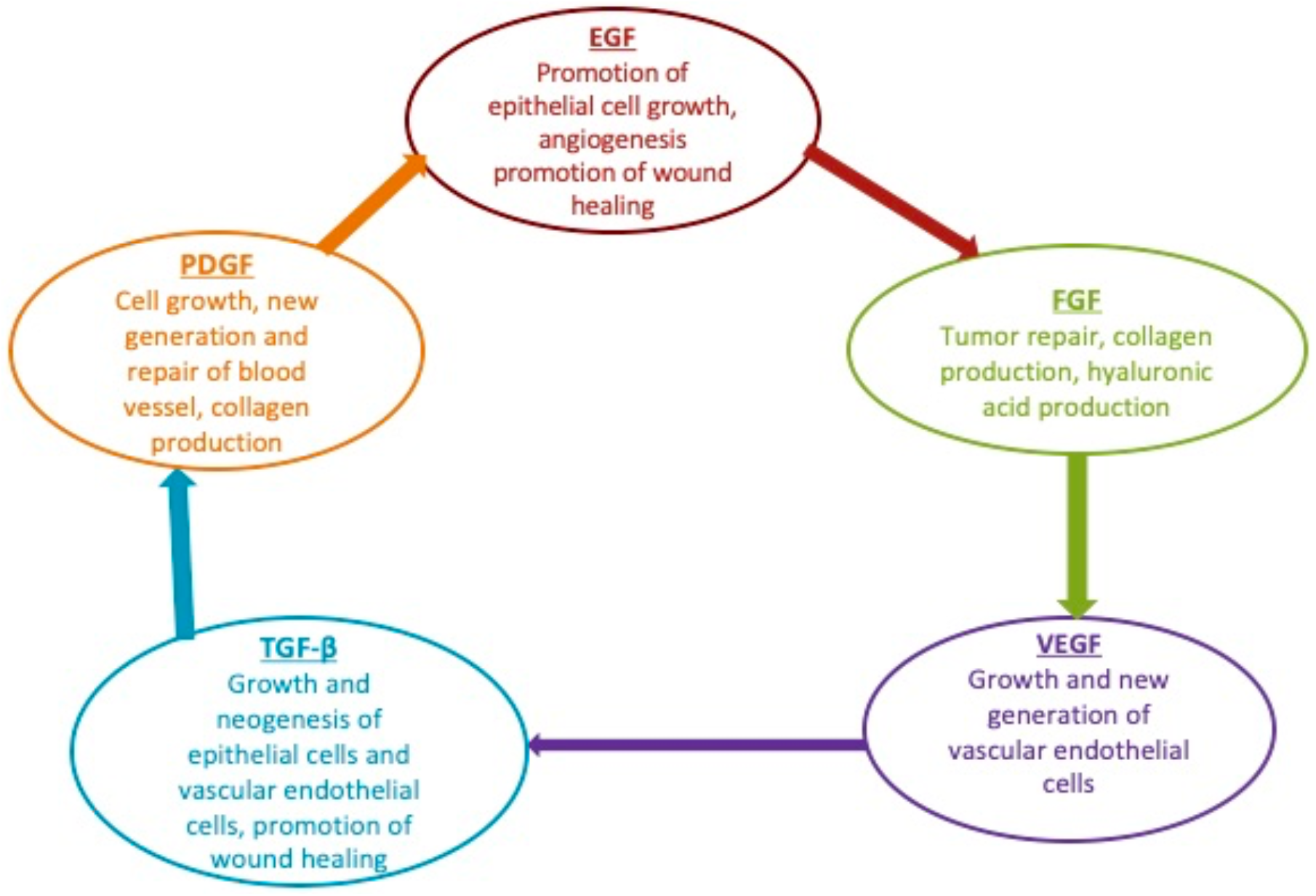
Growth factors typically found in platelet rich plasma (adapted from [39]).

